# An independent external validation of the QRISK3 cardiovascular risk prediction model applied to UK Biobank participants

**DOI:** 10.1101/2022.06.30.22277083

**Authors:** Ruth E. Parsons, Xiaonan Liu, Jennifer A. Collister, David A. Clifton, Benjamin J. Cairns, Lei Clifton

## Abstract

**Background:** The QRISK3 cardiovascular disease (CVD) risk prediction model was derived using primary care data; however, it is frequently used outside of clinical settings. The use of QRISK3 in epidemiological studies without external validation may lead to inaccurate results, however it has been used multiple times on data from UK Biobank. We aimed to externally evaluate the performance of QRISK3 for predicting 10-year risk of cardiovascular events in the UK Biobank cohort.

**Methods:** We used data from the UK Biobank, a large-scale prospective cohort study of 403,370 participants aged 40-69 years recruited between 2006 and 2010 in the United Kingdom (UK). We included participants with no previous history of CVD or statin treatment and the outcome was the first occurrence of coronary heart disease, ischaemic stroke or transient ischaemic attack, derived from linked hospital episode statistics (HES) and death registration data (DRD).

**Results:** Our study population included 233,233 females and 170,137 males, with 9295 and 13,028 incident cardiovascular events, respectively. The overall median follow-up time after recruitment was 11.7 years. The discrimination measure of QRISK3 in the overall population was reasonable (Harrell’s C-Index 0.722 in females and 0.697 in males), this was poorer in older participants (<0.62 in all participants aged 65 or older). QRISK3 had systematic over-prediction of CVD risk in UK Biobank, particularly in older participants, by as much as 20%.

**Conclusions:** QRISK3 had reasonable overall discrimination for the whole study population, which was best in younger participants. The observed CVD risk in UK Biobank participants was lower than that predicted by QRISK3, particularly for older participants. The UK Biobank cohort is known to be healthier than the general population and therefore it is necessary to recalibrate QRISK3 before using it to predict absolute CVD risk in the UK Biobank cohort.

## INTRODUCTION

Cardiovascular diseases (CVDs) are the leading cause of global mortality [1] and healthcare providers need to identify patients at a high risk of CVD to accurately and reliably allocate primary prevention measures. Prognostic models can classify individuals into event risk groups, allowing decisions to be made about their individual health care. There are numerous prediction models designed to estimate the risk of developing CVD in use worldwide, including the Framingham [2], SCORE [3] and QRISK [4,5,6] models. The QRISK models are routinely used in the United Kingdom (UK) by healthcare providers to calculate the ten-year CVD risk of patients during NHS Health Checks [7].

The first QRISK model was published in 2007 with the aim of estimating the ten-year risk of CVD in females and males [4]. This model was derived using the 35 million anonymised health records from practices across the UK that are held in the QResearch primary care database [4]. QRISK was followed by an updated model in 2008 (QRISK2) which included additional risk factors compared to its predecessor [6]. Since 2008, QRISK2 has been updated continually, including extending the age range, adding type 1 diabetes, creating further categories for the smoking variable, and updating the Townsend deprivation score [5,6]. The most up to date QRISK model, QRISK3, was derived in 2017 [6] to incorporate risk factors that were outlined in the National Institute for Health and Care Excellence (NICE) 2014 clinical guideline [8]. The risk factors included in the QRISK3 model can be seen in Box 1.

QRISK3 was validated using data from the Clinical Practice Research Datalink (CPRD) in 2021 and was found to perform well at the overall population level [9]. CPRD contains primary care data which, by definition, has a similar case mix to the QResearch primary care database. The generalisability of a model should be examined in each population that differs in setting to the derivation cohort [10] before the model can be used reliably in that independent population, such as UK Biobank.

Following a literature search we identified multiple studies that have used the QRISK3 scores of UK Biobank participants in their analysis. The extent to which the authors of the identified studies address the discrimination and the calibration of QRISK3 applied to UK Biobank data varies, and the extent to which this may influence their conclusions depends on their study aim. In five of the identified studies, the authors do not address discrimination or calibration [11,12,13,14,15], three only address discrimination [16,17,18] and one quotes a measure of discrimination calculated in another study [19]. The authors of one study found that the mean 10-year QRISK3 predicted CVD risk was greater than the observed 10-year event rate of coronary artery disease (CAD) in UK Biobank [20] and recalibrated QRISK3 for their analyses [20].

These studies are applying a model which was developed and validated on primary care data to a volunteer cohort. The UK Biobank participants tend to be older, to be female, and to live in more socioeconomically affluent areas than non-participants [21]. Additionally, compared to the general population, UK Biobank participants tend to have fewer health conditions and are less likely to be obese, to smoke, and to drink alcohol [21]. The UK Biobank cohort is not representative of the general population of the UK, with evidence of healthy volunteer selection bias [21].

Without considering the generalisability of the QRISK3 model to UK Biobank data, the accuracy of the CVD risk scores used in this context are unknown and therefore study conclusions may be misleading. This paper provides an independent external validation of the QRISK3 model applied to UK Biobank and offers some suggestions for future researchers.

### Box 1

Risk factors included in the QRISK3 model [6]

1. Age at study entry (baseline)
2. Ethnic origin (nine categories)
3. Deprivation (as measured by the Townsend score, where higher values indicate higher levels of material deprivation)
4. Systolic blood pressure
5. Body mass index
6. Total cholesterol: high-density lipoprotein cholesterol ratio
7. Smoking status (non-smoker, former smoker, light smoker (1-9/day), moderate smoker (10-19/day), or heavy smoker (≥20/day))
8. Family history of coronary heart disease in a first degree relative aged less than 60 years
9. Diabetes (type 1, type 2, or no diabetes)
10. Treated hypertension (diagnosis of hypertension and treatment with at least one antihypertensive drug)
11. Rheumatoid arthritis (diagnosis of rheumatoid arthritis, Felty’s syndrome, Caplan’s syndrome, adult onset Still’s disease, or inflammatory polyarthropathy not otherwise specified)
12. Atrial fibrillation (including atrial fibrillation, atrial flutter, and paroxysmal atrial fibrillation)
13. Chronic kidney disease (general practitioner recorded diagnosis of chronic kidney disease stage 3, stage 4 or 5) and major chronic renal disease (including nephrotic syndrome, chronic glomerulonephritis, chronic pyelonephritis, renal dialysis, and renal transplant)
14. Measure of systolic blood pressure variability (standard deviation of repeated measures)
15. Diagnosis of migraine (including classic migraine, atypical migraine, abdominal migraine, cluster headaches, basilar migraine, hemiplegic migraine, and migraine with or without aura)
16. Corticosteroid use (British National Formulary (BNF) chapter 6.3.2 including oral or parenteral prednisolone, betamethasone, cortisone, depo-medrone, dexamethasone, deflazacort, efcortesol, hydrocortisone, methylprednisolone, or triamcinolone)
17. Systemic lupus erythematosus (including diagnosis of SLE, disseminated lupus erythematosus, or Libman-Sacks disease)
18. Second generation “atypical” antipsychotic use (including amisulpride, aripiprazole, clozapine, lurasidone, olanzapine, paliperidone, quetiapine, risperidone, sertindole, or zotepine)
19. Diagnosis of severe mental illness (including psychosis, schizophrenia, or bipolar affective disease)
20. Diagnosis of erectile dysfunction or treatment for erectile dysfunction (BNF chapter 7.4.5 including alprostadil, phosphodiesterase type 5 inhibitors, papaverine, or phentolamine)

## METHODS

### STUDY POPULATION

The study design and methods of the UK Biobank study have been described previously [22,23]. Approximately 9.2 million people were invited by written correspondence to take part in the UK Biobank study, these were individuals aged approximately between 40 to 69 years who were registered with the NHS and lived in a 25-mile radius of one of 22 assessment centres in England, Wales and Scotland [22]. 503,325 individuals attended an assessment centre, which is a response rate of 5.5% [22,23]. The study received ethical approval from the National Health Service’s National Research Ethics Service North West (11/NW/0382). At baseline, all participants provided written informed consent for the study and completed a touch screen questionnaire, a verbal interview and had physical measurements and blood, urine, and saliva samples collected [22,23].

To align with the exclusion criteria used in the derivation of QRISK3 [6], we excluded UK Biobank participants if they had prior diagnosis of CVD, were using prescribed cholesterol lowering medication at cohort entry or had missing Townsend deprivation scores. The participants in UK Biobank (40 to 69 years) are all within the age range required for the QRISK3 model (25 to 84 years).

### DEFINITION OF RISK FACTORS

We matched the risk factors included in the QRISK3 model to the variables available in UK Biobank using a mapping provided by Elliot et al. [16]. When a risk factor required for the QRISK3 model could not be perfectly matched to a UK Biobank field, we used the fields with the closest matches. Details on how each risk factor was defined can be seen in the supplementary material.

### OUTCOMES

The outcome of interest is incident CVD defined in the QRISK3 derivation by a composite outcome of coronary heart disease, ischaemic stroke, or transient ischaemic attack [6]. We derived this outcome using International Classification of Diseases (ICD-9 and ICD-10) and the Office of Population Censuses and Surveys Classification of Interventions and Procedures version 4 codes (OPCS-4) from hospital episode statistics and death registration data. Follow-up time for each participant was calculated as the number of years from date of baseline assessment until the earliest date of the following: CVD event date, death date by other causes, loss to follow-up date or UK Biobank administrative censoring date (England: 2020-11-30; Scotland: 2020-10-31; Wales: 2018-02-28). The supplementary material contains definitions of CVD used in this study according to ICD-9, ICD-10, OPCS-4 and UK Biobank Field IDs (FID).

### STATISTICAL ANALYSIS

As in the derivation of QRISK3, participants with missing Townsend deprivation scores were excluded and those with missing data on ethnicity were assumed to be White [6]. We used the multivariate imputation by chained equations (MICE) package in R to impute missing data on total cholesterol/HDL cholesterol ratio, smoking status, weight, height, SBP and SBP variability, by gender. In the imputation model for males, we included all QRISK3 model predictor risk factors, along with survival outcomes; and for females, we included all gender-specific predictors and survival outcomes but diagnosis of or treatment for erectile dysfunction was removed from the imputation model. Ten imputations were carried out, which is sufficient for high efficiency [24]. We implemented the QRISK3-2017 prediction model using the user-written package ‘QRISK3’ in R [25]. Statistical estimates from the ten imputed datasets were pooled using Rubin’s rules [26] to produce summary estimates and confidence limits, incorporating the additional uncertainty of the imputed datasets.

Model performance of QRISK3 was assessed by both discrimination and calibration. Discrimination measures the ability to distinguish between low and high risk patients [27]; patients with higher risk predictions should have higher event rates than those with lower risk predictions [28,29]. We assessed discrimination overall and in each age group as used in the QRISK3 derivation (35-44, 45-54, 55-64, 65-74 years) at ten years. We used Harrell’s C-Index, a measure which quantifies the correlation between ranked predicted and observed survival [30,31], a C-Index of 0.5 indicates that the risk prediction from the model is no better than chance in predicting patient outcomes and values near 1 indicate that the model is approaching perfect separation of patient outcomes. Additionally, we used Royston and Sauerbrei’s D-Index which measures the amount of variation in risk between individuals with low and high predicted risks [31,32]. The D-Index can be interpreted as the log hazard ratio between the low and high risk groups, with higher values showing greater discrimination and an increase of 0.1 or more over other risk scores is an indicator of improved outcome discrimination [6,31,33]. Additionally, we calculated the R2D statistic overall and in each age group as a measure of explained variation (the proportion to which the model accounts for the dispersion of the data set) tailored towards survival data, based on the D-Index [34]. Higher values of the R^2^_D_ statistic indicate that a higher proportion of the variation in CVD risk in UK Biobank is explained by the dependent covariates included in the QRISK3 model and suggests that there is less residual variation.

Calibration assesses how well the predicted risk corresponds to the observed risk on a group level [28,35]. We assessed calibration graphically by comparing the mean predicted risk with the mean observed risk at ten years, by deciles of the QRISK3 predicted risk distribution. The observed risks were obtained using cumulative incidence Kaplan-Meier estimates at ten years. Calibration was evaluated overall and in each age group.

All analyses were conducted in R (version 4.1.1).

## RESULTS

Data from 502,488 participants in the UK Biobank study were reviewed for eligibility in this study (Figure 1). In line with the QRISK3 derivation exclusion criteria, 623 participants with missing Townsend deprivation scores, a further 90,296 using statins at baseline and finally 8199 with previous diagnosis of CVD were excluded. Median follow-up time was 11.7 years and 92.4% participants had ten years or more of follow-up. Consequently, 233,233 female and 170,137 male participants of the UK Biobank study were included in the analyses. The minimum age of the participants enrolled in UK Biobank was 39.7 for females and 37.4 for males with the maximum being 71.0 and 73.7 respectively. Table 1 shows the baseline characteristics for the included participants and Table 2 shows the quantity of missing data by age and sex. Overall, 18.1% of the data was missing and was replaced by imputation.

**Table 1.** Baseline characteristics of all participants in the UK Biobank cohort and the published QRISK3 derivation cohort [6] by sex. Values are number (percentages) unless otherwise stated.

|  | UK Biobank<br>Female<br>(N=233,233) | QResearch<br>Derivation<br>Female<br>(N=4,019,956) | UK Biobank Male<br>(N=170,137) | QResearch<br>Derivation Male<br>(N=3,869,847) |
| --- | --- | --- | --- | --- |
| **Age (years)** mean (SD) | 56.02 (8.0) | 43.3 (15.3) | 55.78 (8.2) | 42.6 (14.0) |
| **Systolic blood pressure (mmHg)** mean (SD) | 134.40 (19.2) | 123.2 (18.2) | 140.38 (17.4) | 129.2 (16.3) |
| Missing | 897 (0.4) | 692,511 (17.2) | 510 (0.3) | 1,225,165 (31.7) |
| **Measure of systolic blood pressure variability (standard deviation of repeated measures)** mean (SD) | 5.44 (4.5) | 9.3 (6.2) | 5.18 (4.3) | 9.9 (6.8) |
| Missing | 897 (0.4) | 896,135 (22.3) | 510 (0.3) | 1,530,945 (39.6%) |
| **Total cholesterol: high-density lipoprotein cholesterol ratio (mmol/L)** mean (SD) | 3.88 (1.01) | 3.7 (1.2) | 4.62 (1.2) | 4.4 (1.4) |
| Missing | 35,880 (15.4) | 2,421,398 (60.2) | 22,694 (13.3) | 2,402,100 (62) |
| **Family history of coronary heart disease** | 99,896 (42.8) | 481,628 (12.0) | 61,453 (36.1) | 357,987 (9.3) |
| **Self-reported ethnicity** |  |  |  |  |
| White or not stated | 221,300 (94.9) | 3,564,651 (88.7) | 161,300 (94.8) | 3,435,408 (88.8) |
| Indian | 2255 (1.0) | 77,683 (1.9) | 1929 (1.1) | 81,805 (2.1) |
| Pakistani | 538 (0.2) | 39,541 (1.0) | 703 (0.4) | 46,948 (1.2) |
| Bangladeshi | 46 (<0.1) | 31,930 (0.8) | 94 (0.1) | 42,111 (1.1) |
| Other Asian | 670 (0.3) | 53,559 (1.3) | 638 (0.4) | 45,753 (1.2) |
| Black Caribbean | 2337 (1.0) | 37,781 (0.9) | 1293 (0.8) | 30,610 (0.8) |
| Black African | 1380 (0.6) | 77,813 (1.9) | 1325 (0.8) | 71,245 (1.8) |
| Chinese | 879 (0.4) | 33,767 (0.8) | 486 (0.3) | 23,730 (0.6) |
| Other ethnic group | 3828 (1.6) | 103,231 (2.6) | 2369 (1.4) | 92,237 (2.4) |
| **Townsend deprivation score** mean (SD) | -1.43 (3.0) | 0.4 (3.2) | -1.34 (3.10) | 0.5 (3.3) |
| **Body mass index (kg/m<sup>2</sup>)** mean (SD) | 26.69 (5.0) | 25.4 (5.1) | 27.36 (4.02) | 25.9 (4.2) |
| Missing | 1148 (0.5) | 1,093,554 (27.2) | 1098 (0.6) | 1,393,672 (36.0) |
| **Smoking status** |  |  |  |  |
| Non-smoker | 140,470 (60.2) | 2,051,803 (51.0) | 88,708 (52.1) | 1,463,941 (37.8) |
| Ex-smoker | 71,274 (30.6) | 589,521 (14.7) | 59,143 (34.8) | 594,265 (15.4) |
| Light smoker (Less than 10 a day) | 2743 (1.2) | 434,954 (10.8) | 1656 (1.0) | 507,523 (13.1) |
| Moderate smoker (10 to 19 a day) | 7765 (3.3) | 226,128 (5.6) | 5705 (3.4) | 251,170 (6.5) |
| Heavy smoker (20 or over a day) | 4693 (2.0) | 115,890 (2.9) | 5927 (3.5) | 188,857 (4.9) |
| Missing | 6288 (2.7) | 601,660 (15.0) | 8998 (5.3) | 864,091 (22.3) |
| **Atrial fibrillation** | 669 (0.3) | 15,177 (0.4) | 1190 (0.7) | 20,098 (0.5) |
| **Erectile dysfunction** | NA | NA | 698 (0.4) | 90,753 (2.3) |
| **Migraine** | 10,421 (4.5) | 257,825 (6.4) | 2613 (1.5) | 103,995 (2.7) |
| **Rheumatoid arthritis** | 3239 (1.4) | 45,700 (1.1) | 1171 (0.7) | 20,997 (0.5) |
| **Chronic kidney disease** | 227 (0.1) | 19,396 (0.5) | 189 (0.1) | 12,254 (0.3) |
| **Severe mental illness** | 1013 (0.4) | 274,069 (6.8) | 881 (0.5) | 167,115 (4.3) |
| **Systemic lupus erythematosus** | 440 (0.2) | 4010 (0.1) | 45 (<0.1) | 365 (<0.1) |
| **Diabetes type 1** | 17 (<0.1) | 10,060 (0.3) | 16 (<0.1) | 11,617 (0.3) |
| **Diabetes type 2** | 1797 (0.8) | 48,022 (1.2) | 2161 (1.3) | 58,395 (1.5) |
| **Second generation “atypical” antipsychotic use** | 483 (0.2) | 19,140 (0.5) | 456 (0.3) | 20,123 (0.5) |
| **Corticosteroid use** | 1898 (0.8) | 96,955 (2.4) | 1253 (0.7) | 56,533 (1.5) |
| **Treated hypertension** | 29,089 (12.5) | 223,494 (5.6) | 19,167 (11.3) | 164,255 (4.2) |

**Table 2.** Quantity of missing data of all participants in the UK Biobank at baseline by sex and age.

|  | Females |  |  |  | Males |  |  |  |
| --- | --- | --- | --- | --- | --- | --- | --- | --- |
| Age Group (years) | <45<br>(N=27,022) | 45-55<br>(N=74,928) | 55-65<br>(N=96,788) | >65<br>(N=34,495) | <45<br>(N=22,482) | 45-55<br>(N=54,093) | 55-65<br>(N=67,247) | >65<br>(26,313) |
| Systolic blood pressure | 136 (0.5%) | 287<br>(0.38%) | 354<br>(0.37%) | 120 (0.3%) | 80 (0.4%) | 143<br>(0.26%) | 187<br>(0.28%) | 100<br>(0.4%) |
| Measure of systolic blood pressure variability | 136 (0.5%) | 287<br>(0.38%) | 354<br>(0.37%) | 120 (0.3%) | 80 (0.4%) | 143<br>(0.26%) | 187<br>(0.28%) | 100<br>(0.4%) |
| Total cholesterol: high density lipoprotein cholesterol ratio | 4283<br>(15.9%) | 11,794<br>(15.7%) | 14,667<br>(15.2%) | 5136<br>(14.9%) | 3080<br>(13.7%) | 7077<br>(13.1%) | 8970<br>(13.3%) | 3567<br>(13.6%) |
| Body mass index | 151 (0.6%) | 368<br>(0.49%) | 454<br>(0.47%) | 175 (0.5%) | 159 (0.7%) | 352<br>(0.65%) | 422<br>(0.63%) | 165<br>(0.6%) |
| Smoking status | 1098<br>(4.1%) | 2313<br>(3.1%) | 2221<br>(2.3%) | 656 (1.9%) | 1478<br>(6.6%) | 2904<br>(5.4%) | 3337<br>(5.0%) | 1279<br>(4.9%) |

Data are n (%).

**Figure 1.**
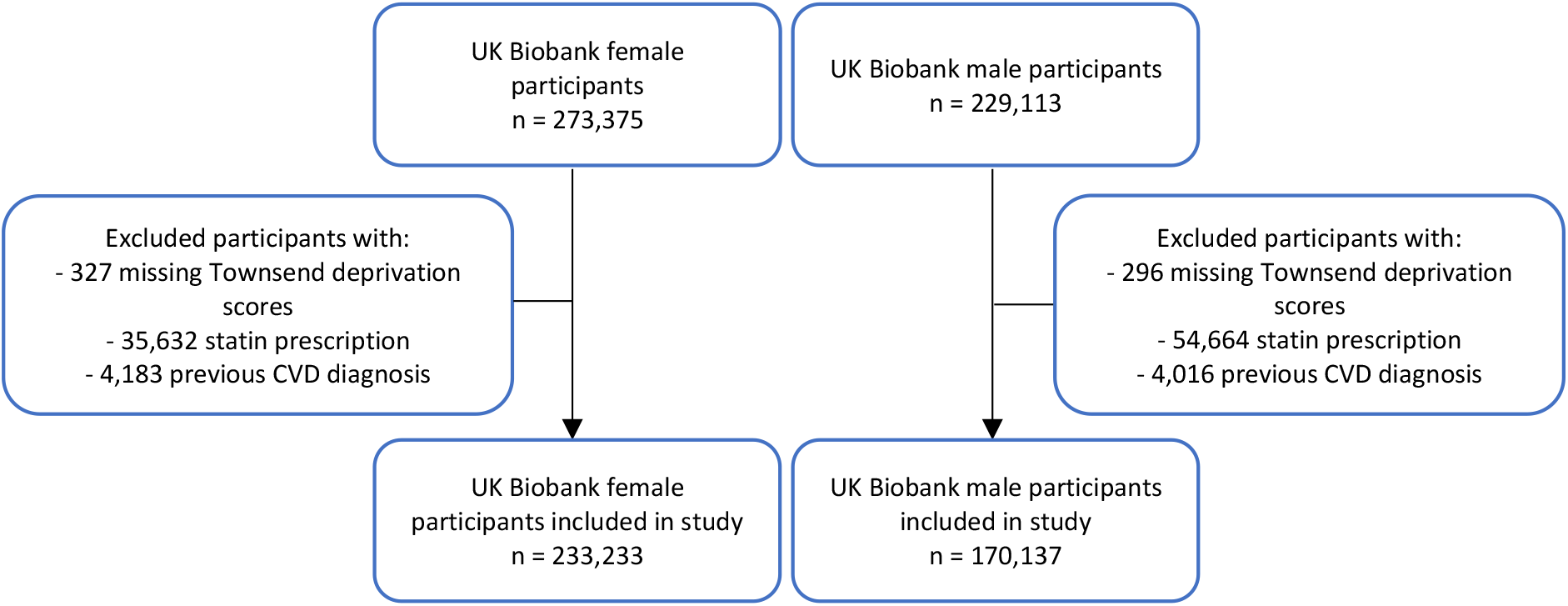
Flow chart of the female and male population used for this study after exclusions for missing Townsend deprivation scores, statin prescription and previous diagnoses of CVD.

The maximum follow-up time in our UK Biobank population (after applying exclusion criteria) is 13.95 years, less than the maximum follow-up time of 15 years in the derivation cohort of QRISK3; therefore, there is no need for us to extrapolate the estimate of this external validation of QRISK3 [28].

### DISCRIMINATION

The QRISK3 scores of the UK Biobank participants shows that there was good overall discrimination for females and reasonable discrimination for males between risk scores as measured by the C-Index and the D-Index (Table 3). As expected, discrimination in this external validation study is not as good as that in the QResearch internal validation study [6] (for females, C-Index: 0.722 for UK Biobank external validation vs 0.880 for interval validation, D-Index: 1.28 vs 2·49; for males, C-Index: 0·697 vs 0·858, D-Index: 1.11 vs 2·26; Table 3).

**Table 3.**
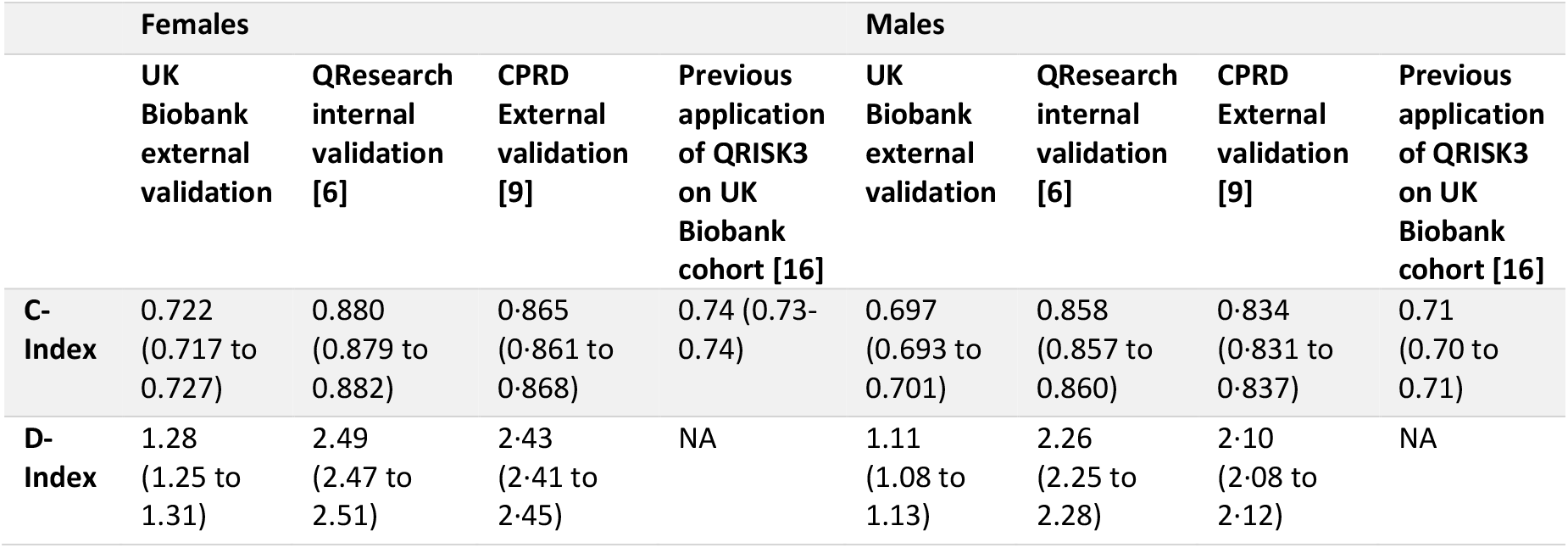
Measures of discrimination performance for QRISK3 in the UK Biobank, QResearch and CPRD cohorts.

|  | Females |  |  |  | Males |  |  |  |
| --- | --- | --- | --- | --- | --- | --- | --- | --- |
|  | UK Biobank external validation | QResearch internal validation [6] | CPRD External validation [9] | Previous application of QRISK3 on UK Biobank cohort [16] | UK Biobank external validation | QResearch internal validation [6] | CPRD External validation [9] | Previous application of QRISK3 on UK Biobank cohort [16] |
| C-Index | 0.722<br>(0.717 to 0.727) | 0.880<br>(0.879 to 0.882) | 0.865<br>(0.861 to 0.868) | 0.74 (0.73-0.74) | 0.697<br>(0.693 to 0.701) | 0.858<br>(0.857 to 0.860) | 0.834<br>(0.831 to 0.837) | 0.71<br>(0.70 to 0.71) |
| D-Index | 1.28<br>(1.25 to 1.31) | 2.49<br>(2.47 to 2.51) | 2.43<br>(2.41 to 2.45) | NA | 1.11<br>(1.08 to 1.13) | 2.26<br>(2.25 to 2.28) | 2.10<br>(2.08 to 2.12) | NA |

Discrimination varied distinctly between age groups, with the best discrimination in the youngest age group (35 to 45 years) and the worst in the oldest age group (65 to 75 years) for both sexes (Table 4). The ability of QRISK3 to discriminate was attenuated as age increased, with the C-Index and D-Index decreasing from 0.722 and 1.39 in the youngest female participants to 0.617 and 0.67 in the oldest female participants. These measures decreased from 0.722 and 1.34 in the youngest male participants to 0.597 and 0.54 in the oldest male participants. Discrimination was better in female participants than male participants overall and by age, with the discriminative ability of QRISK3 in the oldest male participants being scarcely better than a C-Index of 0.5, which would represent chance alone.

**Table 4.** Measures of discrimination performance for QRISK3 in the UK Biobank cohort in each age group, with 95% confidence intervals (CI).

|  | Females |  |  |  | Males |  |  |  |
| --- | --- | --- | --- | --- | --- | --- | --- | --- |
| Age Group (years) | <45 | 45-55 | 55-65 | 65-75 | 35-45 | 45-55 | 55-65 | >65 |
| **C-Index** | 0.722<br>(0.690 to 0.753) | 0.700<br>(0.686 to 0.713) | 0.657<br>(0.649 to 0.665) | 0.617<br>(0.606 to 0.627) | 0.725<br>(0.701 to 0.748) | 0.673<br>(0.663 to 0.683) | 0.631<br>(0.625 to 0.638) | 0.597<br>(0.588 to 0.606) |
| **D-Index** | 1.39<br>(1.20 to 1.58) | 1.20<br>(1.12 to 1.28) | 0.92<br>(0.87 to 0.97) | 0.67<br>(0.61 to 0.73) | 1.34<br>(1.20 to 1.48) | 1.01<br>(0.95 to 1.07) | 0.75<br>(0.71 to 0.79) | 0.54<br>(0.49 to 0.60) |

### CALIBRATION

Figure 2 shows the agreement between the ten-year observed and QRISK3 predicted CVD risk, grouped according to the decile of their respective QRISK3 score, such that the 10% of patients with the lowest QRISK3 score were binned into the first decile and so forth. The predicted probability within each decile group is plotted as the blue squares in Figure 2 and was calculated as the average QRISK3 score within that group. The observed 10-year CVD probability within each group is plotted as the orange circles in Figure 2 and was calculated using the Kaplan-Meier method (to account for right censoring). The plot suggests that QRISK3 systematically over-predicts the probability of CVD for UK Biobank participants, with the magnitude of overprediction increasing at higher risk deciles.

**Figure 2.**
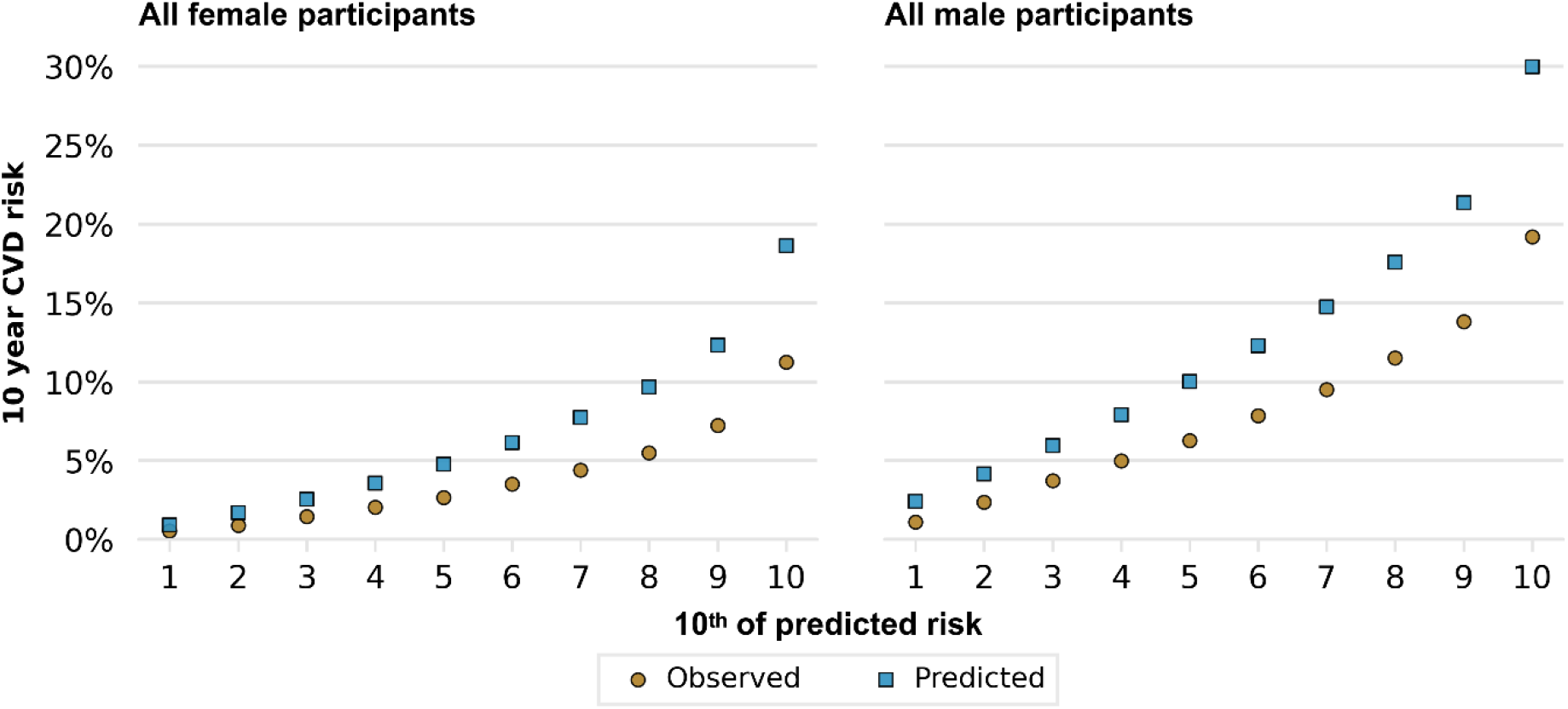
Calibration of QRISK3 at ten years for female and male participants of UK Biobank overall. The cumulative Kaplan-Meier observed CVD probability in each tenth of risk is denoted by the orange circular markers and the mean predicted QRISK3 score in each tenth of risk is denoted by the blue square markers.

Figure 3 shows the agreement between the ten-year observed and QRISK3 predicted CVD risk, grouped according to the decile of the participant’s respective QRISK3 score by age group and can be interpreted in the same way as Figure 2. Figure 3 suggests that the overprediction of CVD for UK Biobank participants seen in Figure 2 may be driven by the increasing magnitude of overprediction for older participants.

**Figure 3.**
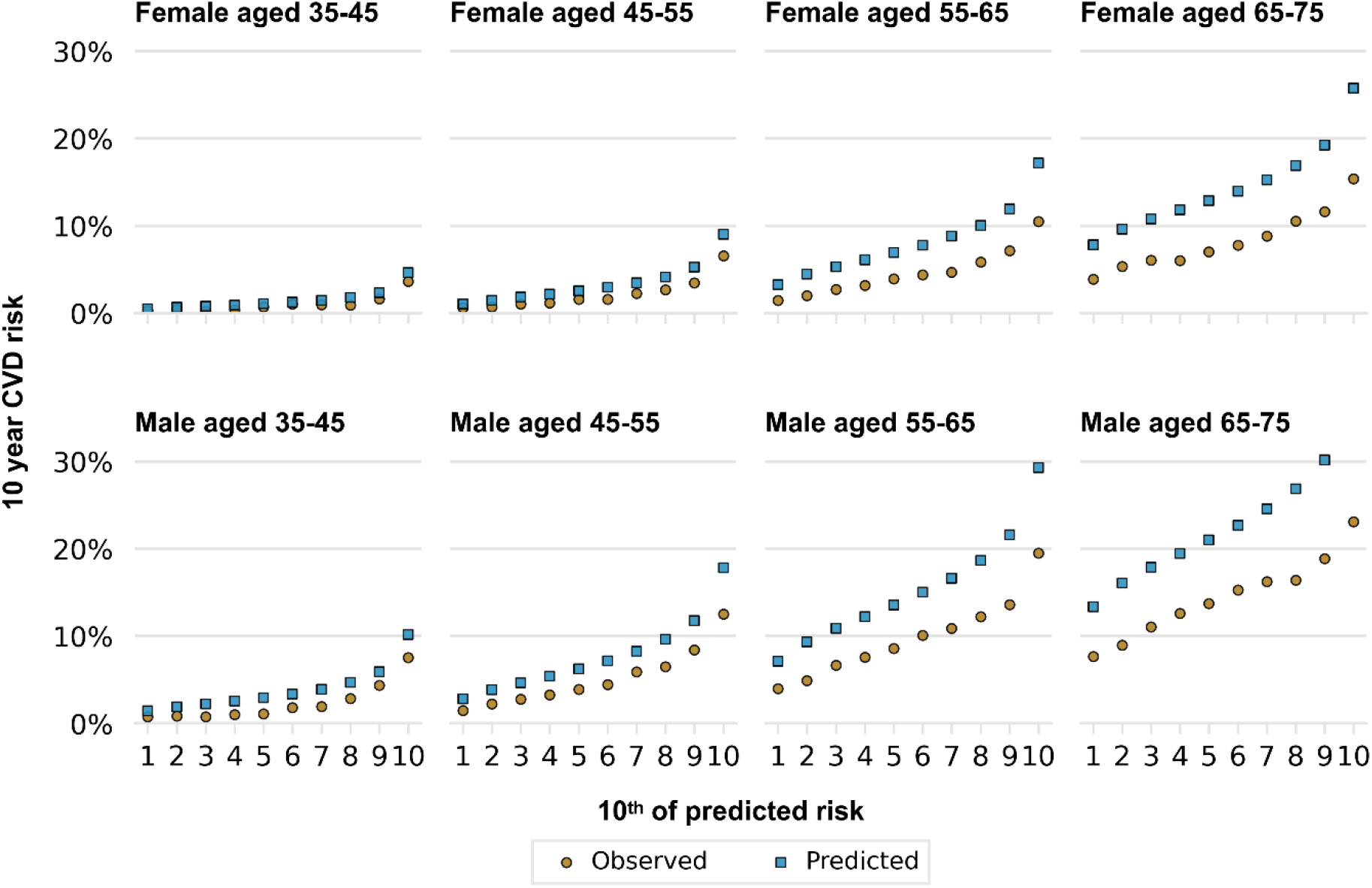
Calibration of QRISK3 at ten years for female and male participants of UK Biobank in each age group. The cumulative Kaplan-Meier observed CVD probability in each tenth of risk is denoted by the orange circular markers and the mean predicted QRISK3 score in each tenth of risk is denoted by the blue square markers.

### OVERALL MODEL PERFORMANCE

Table 5 displays explained variation for QRISK3 model in the UK Biobank cohort overall and in age groups, using the R^2^_D_ measure described by Royston and Sauerbrei [32]. Overall, for female participants QRISK3 explained 28.2% of the variation in time to a cardiovascular outcome, this was 22.6% for males. This contrasted with an R2D of 59.6% for female patients and 55.0% for male patients in the QResearch internal validation [6]. The overall model performance decreases with age, with the best performance seen in the youngest age group (35 to 45 years) and the worst in the oldest age group (65 to 75 years) for both sexes.

**Table 5.** R^2^_D_ measure of explained variation for the UK Biobank external validation of QRISK3 Model in the UK Biobank cohort overall and for each age group, as well as overall for the QResearch internal validation.

| Age Group (years) | Female |  |  |  |  | Males |  |  |  |  |
| --- | --- | --- | --- | --- | --- | --- | --- | --- | --- | --- |
|  | All | <45 | 45-55 | 55-65 | >65 | All | <45 | 45-55 | 55-65 | >65 |
| UK Biobank External validation | 28.2% | 31.5% | 25.5% | 16.7% | 9.7% | 22.6% | 30.1% | 19.2% | 11.8% | 6.6% |
| Internal validation [6] | 59.6% | NA | NA | NA | NA | 55.0% | NA | NA | NA | NA |
| NA = not applicable as these measures were not reported in the QResearch internal validation. |  |  |  |  |  |  |  |  |  |  |

## DISCUSSION

### PRINCIPAL FINDINGS

In this external validation, QRISK3 had good overall discrimination for females (C-Index: 0.722, D-Index: 1.28) and reasonable discrimination for males (C: 0.697, D: 1.11) in UK Biobank. The best discriminative accuracy of QRISK3 was seen in the youngest age group for both sexes (35 to 45 years; C: 0.722, D: 1.39 for females and C: 0.725, D: 1.34 for males), but this discriminative accuracy diminished with age.

The good overall discrimination of QRISK3 in UK Biobank is consistent with studies that have applied the model to this population previously [11,16,17,18]. This good overall discrimination is the result of the good discrimination in younger participants in UK Biobank participants, a finding which is mirrored in Riveros-McKay et al. [18]. Discrimination statistics by age group were not reported in the QResearch internal validation study [6], preventing any comparison of the age gradient finding from this study. However, these findings are consistent in direction and magnitude with the CPRD external validation study [9]. There is no marked difference between the quantity of missing values in each age group in the UK Biobank for the risk factors included in QRISK3.

Calibration was generally poor for QRISK3 in UK Biobank, with over-prediction of cardiovascular events for both sexes and in all age groups at ten years. The overall model performance measure R^2^_D_ was 28.2% for female participants and 22.6% for male participants. As with the measures of discrimination in this study, the overall model performance masked the variation in R^2^_D_ for participants of different ages.

### INTERPRETATION OF FINDINGS

This study is the first to investigate the calibration of QRISK3 in UK Biobank. Our results contrasted with findings from the QResearch internal validation cohort [6] and CPRD external validation cohort [9], where calibration was very good overall. Such inconsistency between studies is likely the result of the difference between the UK Biobank cohort and the QResearch and CPRD cohorts, which were derived from primary care databases. UK Biobank is known not to be representative of the general UK population, with participants tending to be older, female, live in less socioeconomically deprived areas, be less obese, smoke less, drink less alcohol and have fewer health conditions and there is evidence of healthy volunteer bias [21].

The calibration of QRISK3 was less good in older UK Biobank participants, with the greatest over-prediction of actual CVD risk for the oldest age group in both sexes. Although it was not the case in the QResearch internal validation study [6], poor calibration in older participants was also found in the CPRD external validation cohort [9]. This may reflect the CPRD cohort being slightly older than the QResearch cohort on average, and the UK Biobank cohort being considerably older than both.

Problems associated with poorly calibrated risk prediction models have been explored more in clinical settings than for epidemiological studies, where they may lead to spurious findings [36]. For example, studies that compare risk estimates between cohort groups using a poorly calibrated model are unlikely to be comparing true risks [36]. The research objective of a study defines the amount of consideration that should be given minimising inaccuracy of results to the performance of a model [36].

There are strategies for minimising inaccuracy of results from a model and to improve model performance when applying a risk prediction model to an external dataset. These include model recalibration, considering using an alternative model, and collecting the most appropriate risk factors in cohort studies; these options are discussed further by Parsons, et al. [36].

### STRENGTHS AND LIMITATIONS

The major strength of this study is the large sample size, with 406,616 participants of UK Biobank included, and 22,323 incident cardiovascular outcomes. This study also has high completeness of the outcome, with only 1298 (<0.01%) participants lost to follow-up, whereas the CPRD external validation study of QRISK3 reported a large loss to follow-up (almost two thirds of both men and women were censored due to deregistration or to having less than 10 years of follow-up before the end of the study) [9].

This study followed the transparent reporting of a multivariable prediction model for individual prognosis or diagnosis (TRIPOD) statement [37], covering all 22 checklist items that are essential for good reporting of studies that validate multivariable prediction models. We have performed a thorough external validation of QRISK3, compared to many studies that have reported some form of external validation [38].

One limitation of this study is the accuracy of coding of the variables from UK Biobank data fields to the risk factors required in QRISK3, with several assumptions made about the data. However, this study is important in assessing QRISK3 in a population that is different from a primary care cohort, as QRISK3 has previously been applied outside of clinical settings, including in six UK Biobank studies previously [11,12,15,16,17,18].

There are likely effects of the prediction paradox [39,40], as well as the previous QRISK3 internal [6] and external [9] validations, where the behaviour of the individuals in the cohorts is likely to have been influenced by the predictions of the previous QRISK models used in their medical care. This may invalidate the QRISK3 predictions.

## CONCLUSION

QRISK3 over-predicts CVD risk for participants of UK Biobank, with the magnitude of this over prediction increasing by age. QRISK3 has reasonable overall discrimination for UK Biobank participants, however the discriminative accuracy of the model declines for older participants. Noting the differences in case-mix between UK Biobank and primary care data, researchers using UK Biobank data that require a CVD risk prediction model that is well calibrated or has good discriminatory prediction for older participants may want to consider recalibrating QRISK3 or using an alternative model.

## Supporting information

TRIPOD Statement

## Data Availability

UK Biobank data are available through a procedure described at http://www.ukbiobank.ac.uk/using-the-resource/.

## Acknowledgements

We thank the participants of the UK Biobank and the study team for enabling us to conduct this research.

