## Supplementary Material for "An independent external validation of the QRISK3 cardiovascular risk prediction model applied to UK Biobank participants"

S.1 DEFINITION OF VARIABLES FOR QRISK3

This section outlines the UK Biobank FIDs and linked records that we have used for each of the QRISK3 variables, along with any assumptions made. In cases where variables required in QRISK3 had non-perfect UK Biobank fields matches, we used the fields with the closest matches.

**Diagnosis of disease variables**

We used either self-reported non-cancer illness at baseline (FID 20002) or linked Hospital Episode Statistics (HES) data prior to baseline to derive the diagnosis of rheumatoid arthritis, diabetes (type 1 and 2), systemic lupus erythematosus, atrial fibrillation, chronic kidney disease, migraine, severe mental illness, and erectile dysfunction variables. Disease status was obtained according to International Classification of Disease (ICD) -9 and -10 codes for the HES data, as in the derivation of QRISK3 [6] and demonstrated in a previous mapping of UK Biobank variables to QRISK3 [17], these can be seen in Table S.1.

**Table S.1**
Definitions of diagnosis of disease variables for QRISK3 according to ICD-9, ICD-10, OPCS-4 and UK Biobank FIDs.

| Variable | Self-Reported Data Field Code (FID 20002) | ICD9, ICD10 and OPSC-4 Codes | Treatment and Medication Data Field Code (FID 20003) | Other data fields |
|---|---|---|---|---|
| **Type 1 Diabetes** | 1222 | ICD-10 E10, O240; ICD9 25001, 25011, 25021, 25031, 25041, 25051, 25061, 25071, 25081, 25091, 25003, 25013, 25023, 25033, 25043, 25053, 25063, 25073, 25083, 25093 | | |
| **Type 2 Diabetes** | 1223,1220 | ICD-10 E11, O241; ICD-9 25000, 25010, 25020, 25030, 25040, 25050, 25060, 25070, 25080, 25090, | 1140868902, 1140874646, 1140874674, 1140874718, 1140874744, 1140883066, 1140884600, 1141152590, 1141157284, 1141168660, 1141171646, 1141173882, 1141189090 | FID 2443 = 1, FID 30750 ≥ 48 (HbA1c measurement ≥ 48 mmol/mol) |

| | | |
|---|---|---|
| | | 25002, 25012, 25022, 25032, 25042, 25052, 25062, 25072, 25082, 25092 |
| **Rheumatoid Arthritis** Including rheumatoid arthritis, Felty's syndrome, Caplan's syndrome, adult onset Still's disease, or inflammatory polyarthropathy not otherwise specified | 1464 | ICD-10 M05, M06; ICD-9 714 |
| **Atrial Fibrillation** Including atrial fibrillation, atrial flutter, and paroxysmal atrial fibrillation | 1471, 1483 | ICD-10 I48; ICD-9 4273 and 4270; OPCS-4 K622 and K623 |
| **Chronic Kidney Disease** Including chronic kidney disease stages 3, 4 or 5 and major chronic renal disease; nephrotic syndrome, chronic glomerulonephritis, chronic pyelonephritis, renal dialysis, and renal transplant), | 1192, 1519, 1609 | ICD-10 N183, N184, N185; ICD-9 5853, 5855, 5810, 5820, 5900, V420, V451 |
| **Migraine** Including classic migraine, atypical migraine, abdominal migraine, cluster headaches, basilar migraine, hemiplegic migraine, and migraine with or without aura | 1265 | ICD-10 G43, G440, N943; ICD-9 346 |
| **Systemic Lupus Erythematosus** Including diagnosis of systemic lupus erthematosus, disseminated lupus erythematosus, or Libman-Sacks disease | 1381 | ICD-10 M32; ICD-9 7100 |
| **Severe Mental Illness** Including psychosis, schizophrenia, or bipolar affective disease | 1289, 1291 | ICD-10 F03, F068, F09, F20, F22, F23, F259, F28, F29, F31, F39, F53, F333; ICD9 295, 298, 296 |

| | | | |
|---|---|---|---|
| **Erectile Dysfunction** Including treatment for erectile dysfunction (BNF chapter 7.4.5 including alprostadil, phosphodiesterase type 5 inhibitors, papaverine, or phentolamine | 1518 | ICD-10 N484; ICD-9 60784 | 1141168936, 1141168948, 1141168944, 1141168946, 1140869100, 1140883010 |

We additionally used self-reported treatment and medication reported to clinic nurses at baseline (FID 20003) to determine a diagnosis of diabetes type 2 and erectile dysfunction, under the assumption that those who take related medication have a diagnosis of the disease. Additionally, a reported diagnosis of diabetes by a doctor (FID 2443) with a measurement of glycated haemoglobin (HbA1c) greater than or equal to 48 mmol/mol (FID 30750) was seen to indicate a diagnosis of diabetes type 2, as was done in a previous mapping [17].

We coded diagnosis of each disease as a binary variable, as in the original QRISK3 model [6], with 1 indicating the diagnosis of the disease and 0 indicating no diagnosis of the disease.

**Treatment and medication variables**

We ascertained the use of antihypertensives, corticosteroids and atypical antipsychotics from FID 20003, self-reported treatment and medication reported to clinic nurses at baseline, which excluded short-term medications or prescribed medication that was not taken. We additionally used FIDs 6177 and 6152, the regular use of antihypertensives for males and females respectively. Table S.2 contains definitions of the treatment and medication variables according to the coding of UK Biobank FIDs.

The QRISK3 derivation [6] defines the use of treatments and medications at baseline as being from at least two prescriptions, with the most recent one no more than 28 days before baseline. It is not possible to ascertain this level of information from the fields in UK Biobank. For this reason, we have used the information that is available in UK Biobank FID 20003, 6177 and 6152 under the assumption that they meet this definition.

We coded use of each treatment or medication in QRISK3 as a binary variable, as in the original QRISK3 model [6], with 1 indicating the use of the treatment or medication and 0 indicating no use of the treatment or medication.

**Table S.2**
Definitions of treatment and medication variables for QRISK3 according to ICD-9, ICD-10, OPCS-4 and UK Biobank FIDs.

| Variable | Self-Reported Data Field Code (FID 20002) | ICD9, ICD10 and OPSC-4 Codes | Treatment and Medication Data Field Code (FID 20003) | Other data fields |
|---|---|---|---|---|
| **Treated Hypertension** Diagnosis of hypertension and treatment with at least one antihypertensive drug | | | 1140860192, 1140860292, 1140860696, 1140860728, 1140860750, 1140860806, 1140860882, 1140860904, 1140861088, 1140861190, 1140861276, 1140866072, 1140866078, 1140866090, 1140866102, 1140866108, 1140866122, 1140866138, 1140866156, 1140866162, 1140866724, 1140866738, 1140868618, 1140872568, 1140874706, 1140874744, 1140875808, 1140879758, 1140879760, 1140879762, 1140879802, 1140879806, 1140879810, 1140879818, 1140879822, 1140879826, 1140879830, 1140879834, 1140879842, 1140879866, 1140884298, 1140888552, 1140888556, 1140888560, 1140888646, 1140909706, 1140910442, 1140910614, 1140916356, 1140923272, 1140923336, 1140923404, 1140923712, 1140926778, 1140928226, 1141145660, 1141146126, 1141152998, 1141153026, 1141164276, 1141165470, 1141166006, 1141169516, 1141171336, 1141180592, 1141180772, 1141180778, 1141184722, 1141193282, 1141194794, 1141194810 | FID 6177 = “Blood Pressure Medication” or FID 6152 = “Blood Pressure Medication” |
| **Corticosteroid Use** British National Formulary (BNF) chapter | | | 1140874790, 1140874816, 1140874896.00, 1140874930, | |

| | | | |
|---|---|---|---|
| 6.3.2 including oral or parenteral prednisolone, betamethasone, cortisone, depo-medrone, dexamethasone, deflazacort, efcortesol, hydrocortisone, methylprednisolone, or triamcinolone | | | 1140874976, 1141145782, 1141173346 |
| **Second Generation 'atypical' Antipsychotic Use** Including amisulpride, aripiprazole, clozapine, lurasidone, olanzapine, paliperidone, quetiapine, risperidone, sertindole, or zotepine | | | 1140867420, 1140867444, 1140927956, 1140928916, 1141152848, 1141153490, 1141169714, 1141195974 |

**Sociodemographic and lifestyle variables**

Other than diagnoses of disease and treatment and medication status, additional sociodemographic and lifestyle variables required for QRISK3 (Box 1) were derived using a range of fields from the UK Biobank study. QRISK3 requires some of these variables to be coded as binary, some as categorical and some as continuous, the nature of these variables can be seen in Table S.3, along with the levels for categorical variables.

We derived the QRISK3 variables; ethnic origin, Townsend deprivation scores and sex from the exact matches of these variables in UK Biobank; FID 21000, FID 189 and FID 31. Townsend deprivation score was calculated immediately prior to the participant joining the UK Biobank cohort, based on place of residence and the census.

We generated BMI by dividing UK Biobank FID 50, weight at baseline, by the square of standing height at baseline (FID 21002) converted to meters. We derived the age of participants in years by calculating the time between the year of birth (FID 34), month of birth (FID 52), and the date that the individual attended the baseline assessment centre (FID 53).

The UK Biobank study collected data on smoking habits of participants using multiple variables, for this reason it was not possible to derive a variable with the exact levels used in QRISK3. We obtained the levels of smoking status by assuming that individuals reporting their current smoking as 'only occasionally' at baseline (FID 1239) were light smokers, individuals reporting their smoking status as 'never' (FID 20116) were non-smokers, and individuals reporting their smoking status as 'previous' (FID 20116) were former smokers. For individuals reporting that they currently smoked in FID 1239 or 20116, the number of cigarettes smoked daily (FID 3456) was used to derive their level of smoking.

We ascertained systolic blood pressure (SBP) using the mean of the automated (FID 4080) or manual (FID 93) readings at the initial assessment visit and the first repeat assessment visit, as has been done in a previous study [17]. There is no field in UK Biobank for variability in SBP, we therefore derived this variable as the standard deviation between two automated or manual SBP readings at baseline (FID 4080 and 93).

Total serum cholesterol and high-density lipoprotein (HDL) cholesterol were derived from enzymatic assays collected at baseline of the UK Biobank study and reported in FID 30690 and FID 30760, respectively. We subsequently calculated the ratio of these measurements to obtain the ratio measure required in QRISK3.

The QRISK3 derivation [6] defines family history of coronary heart disease (CHD) as CHD in a first degree relative aged less than 60 years, however, it is not possible to obtain this level of information from the fields of UK Biobank. We therefore used the UK Biobank fields illnesses in father (FID 20107), illnesses in mother (FID 20110), and illnesses of siblings (FID 20111), under the assumption that the level 'heart disease' of these illnesses was CHD and that the relative was aged less than 60 years at diagnosis.

**Table S.3**
Coding of the levels of the sociodemographic and lifestyle variables in QRISK3 **[18]**.

| Risk Factor | QRISK3 variable |
|---|---|
| **Ethnic origin** | Categorical with levels:<br>1 White or not stated<br>2 Indian<br>3 Pakistani<br>4 Bangladeshi<br>5 Other Asian<br>6 Black Caribbean<br>7 Black African<br>8 Chinese<br>9 Other ethnic group |
| **Townsend deprivation score** | Continuous |
| **Body mass index** | Continuous |
| **Smoking** | Categorical with levels:<br>1 non-smoker<br>2 ex-smoker<br>3 light smoker (less than 10)<br>4 moderate smoker (10 to 19)<br>5 heavy smoker (20 or over) |
| **Age** | Continuous |
| **Sex** | Binary:<br>0 male<br>1 female |
| **Systolic blood pressure** | Continuous |
| **Systolic blood pressure variability** | Continuous |
| **Total cholesterol to HDL ratio** | Continuous |
| **Family history of coronary heart disease (CHD) in first degree relative aged less than 60 years** | Binary:<br>0 no<br>1 yes |

**Outcome**

The outcome of interest is CVD defined in the original QRISK3 derivation by a diagnosis of coronary heart disease, ischaemic stroke or transient ischaemic attack [6]. We derived this outcome from the self-reporting of CVD at any visit (FID 20002), self-reported CVD related operations (FID 20004) or vascular/heart problems diagnosed by a doctor (FID 6150) after baseline. Linked HES and operative procedures (using the Office of Population Censuses and Surveys (OPCS) 4 codes) for CVD were also included. Table S.4 contains definitions of CVD used in this study according to ICD-9, ICD-10, OPCS-4 and UK Biobank FIDs.

We coded the outcome as a binary variable, with 1 indicating a participant who experiences an outcome and 0 indicating a participant who does not experience an outcome.

**Table S.4**
Coding for cardiovascular disease for QRISK3 according to ICD-9, ICD-10, OPCS-4 and UK Biobank FIDs.

| Data source | ICD-10 | ICD-9 | OPCS-4 | Non-Cancer Illness Code (FID 20002) | Operation Code (FID 20004) | Vascular/heart problems diagnosed by a doctor (FID 6150) |
|---|---|---|---|---|---|---|
| **Codes** | G45, I20, I21, I22, I23, I24, I25, I63, I64 | 410, 411, 412, 413, 414, 434, 436 | K40, K41, K42, K43, K44, K45, K46, K47.1, K49, K50, K75 | 1074, 1075, 1082, 1583 | 1070, 1071. 1095, 1105, 1109, 1514 | 1, 2, 3 |
