## Supplementary material for "An independent external validation of the QRISK3 cardiovascular risk prediction model applied to UK Biobank participants": TRIPOD Statement

### TRIPOD Checklist: Prediction Model Validation

| Section/Topic | Item | Checklist Item | Page |
|---|---|---|---|
| **Title and abstract** | | | |
| Title | 1 | Identify the study as developing and/or validating a multivariable prediction model, the target population, and the outcome to be predicted. | 1 |
| Abstract | 2 | Provide a summary of objectives, study design, setting, participants, sample size, predictors, outcome, statistical analysis, results, and conclusions. | 3 |
| **Introduction** | | | |
| Background and objectives | 3a | Explain the medical context (including whether diagnostic or prognostic) and rationale for developing or validating the multivariable prediction model, including references to existing models. | 3 |
| | 3b | Specify the objectives, including whether the study describes the development or validation of the model or both. | 4 |
| **Methods** | | | |
| Source of data | 4a | Describe the study design or source of data (e.g., randomized trial, cohort, or registry data), separately for the development and validation data sets, if applicable. | 8 |
| | 4b | Specify the key study dates, including start of accrual; end of accrual; and, if applicable, end of follow-up. | 8 |
| Participants | 5a | Specify key elements of the study setting (e.g., primary care, secondary care, general population) including number and location of centres. | 8 |
| | 5b | Describe eligibility criteria for participants. | 8 |
| | 5c | Give details of treatments received, if relevant. | NA |
| Outcome | 6a | Clearly define the outcome that is predicted by the prediction model, including how and when assessed. | 8/9 |
| | 6b | Report any actions to blind assessment of the outcome to be predicted. | NA |
| Predictors | 7a | Clearly define all predictors used in developing or validating the multivariable prediction model, including how and when they were measured. | Supplement |
| | 7b | Report any actions to blind assessment of predictors for the outcome and other predictors. | NA |
| Sample size | 8 | Explain how the study size was arrived at. | 8 |
| Missing data | 9 | Describe how missing data were handled (e.g., complete-case analysis, single imputation, multiple imputation) with details of any imputation method. | 9 |
| Statistical analysis methods | 10c | For validation, describe how the predictions were calculated. | Supplement |
| | 10d | Specify all measures used to assess model performance and, if relevant, to compare multiple models. | 9/10 |
| | 10e | Describe any model updating (e.g., recalibration) arising from the validation, if done. | NA |
| Risk groups | 11 | Provide details on how risk groups were created, if done. | NA |
| Development vs. validation | 12 | For validation, identify any differences from the development data in setting, eligibility criteria, outcome, and predictors. | 12/18/19/20 |
| **Results** | | | |
| Participants | 13a | Describe the flow of participants through the study, including the number of participants with and without the outcome and, if applicable, a summary of the follow-up time. A diagram may be helpful. | 11 |
| | 13b | Describe the characteristics of the participants (basic demographics, clinical features, available predictors), including the number of participants with missing data for predictors and outcome. | 12 |
| | 13c | For validation, show a comparison with the development data of the distribution of important variables (demographics, predictors and outcome). | 12 |
| Model performance | 16 | Report performance measures (with CIs) for the prediction model. | 13-17 |
| Model-updating | 17 | If done, report the results from any model updating (i.e., model specification, model performance). | NA |
| **Discussion** | | | |
| Limitations | 18 | Discuss any limitations of the study (such as nonrepresentative sample, few events per predictor, missing data). | 19/20 |
| Interpretation | 19a | For validation, discuss the results with reference to performance in the development data, and any other validation data. | 18/19 |
| | 19b | Give an overall interpretation of the results, considering objectives, limitations, results from similar studies, and other relevant evidence. | 18/19 |
| Implications | 20 | Discuss the potential clinical use of the model and implications for future research. | 20 |
| **Other information** | | | |
| Supplementary information | 21 | Provide information about the availability of supplementary resources, such as study protocol, Web calculator, and data sets. | Throughout |
| Funding | 22 | Give the source of funding and the role of the funders for the present study. | 1 |

### TRIPOD Checklist: Prediction Model Validation

We recommend using the TRIPOD Checklist in conjunction with the TRIPOD Explanation and Elaboration document.
